# RISK FACTORS FOR THE DEVELOPMENT OF HOSPITAL-ACQUIRED PEDIATRIC VENOUS THROMBOEMBOLISM: DEALING WITH POTENTIALLY CAUSAL AND CONFOUNDING RISK FACTORS USING DIRECTED ACYCLIC GRAPH (DAG) ANALYSIS

**DOI:** 10.1101/2020.07.17.20153718

**Authors:** Leonardo R. Campos, Maurício Petroli, Flavio R. Sztajnzbok, Elaine S. Costa, Leonardo R. Brandão, Marcelo G. P. Land

## Abstract

**Introduction:** Hospital-acquired venous thromboembolism (HA-VTE) in children comprises multiple risk factors that should not be evaluated separately due to collinearity and multiple cause and effect relationships. This is one of the first case-control study of pediatric HA-VTE risk factors using Directed Acyclic Graph (DAG) analysis.

**Material and Methods:** Retrospective, case-control study with 22 cases of radiologically proved HA-VTE and 76 controls matched by age, sex, unit of admission, and period of hospitalization. Descriptive statistics was used to define distributions of continuous variables, frequencies, and proportions of categorical variables, with a comparison between cases and controls. Due to many potential risk factors of HA-VTE, a directed acyclic graph (DAG) model was created to identify confounding, reduce bias, and increase precision on the analysis. The final model consisted of a DAG-based conditional logistic regression. The study was approved by the Institutional Review Board (CAAE 58056516.0.0000.5264).

**Results:** In the initial univariable model, the following variables were selected as potential risk factors for HA-VTE: length of stay (LOS, days), ICU admission in the last 30 days, LOS in ICU, infection, central venous catheter (CVC), L-asparaginase, heart failure, liver failure and nephrotic syndrome. The final model (table 1) revealed LOS (OR=1.108, 95%CI=1.024-1.199, p=0.011), L-asparaginase (OR=27.184, 95%CI=1.639-450.982, p=0.021), and nephrotic syndrome (OR=31.481, 95%CI=1.182-838.706, p=0.039) as independent risk factors for HA-VTE.

**Conclusion:** The DAG-based approach was useful to clarify the influence of confounders and multiple causalities of HA-VTE. Interestingly, CVC placement - a known thrombotic risk factor highlighted in several studies - was considered a confounder, while LOS, L-asparaginase use and nephrotic syndrome were confirmed as risk factors to HA-VTE. Large confidence intervals are related to the sample size, however the results were significant.

**Highlights:**

1. HA-VTE comprises multiple risk factors that should not be evaluated separately due to collinearity and confounding
2. Directed Acyclic Graph (DAG) helps to clarify collinearity and confounding related to multiple cause and effect relationships that exist in HA-VTE risk factors
3. This is a novel study using DAG-based logistic regression to evaluate risk factors for HA-VTE in children
4. We reported the importance of medical conditions on the genesis of HA-VTE using a DAG-based approach, which makes it possible to clarify the influence of confounders and multiple causalities, such as catheter, a significant risk factor highlighted in several studies

## Introduction

In the past two decades, venous thromboembolism (VTE) has been recognized as an essential complication in pediatric patients. In the last ten years, VTE incidence has reached 42-58 cases of venous thromboembolism (VTE) for every 100,000 children admitted, according to information from the database of pediatric patients admitted to hospitals in the United States of America (USA). This same database showed that the incidence of VTE is much higher in hospitalized children (40.2-10,000 versus 7.8/10,000 outpatients). Deep venous thrombosis (DVT) is currently considered the second leading cause of preventable harm in hospitalized patients, according to a study conducted in 80 pediatric hospitals in the USA(1–3). The clinical consequences of DVT in children are significant since it is estimated that around 25% of children with DVT in the extremities develop signs and/or symptoms of chronic venous insufficiency (known as post-thrombotic syndrome)(4) and 16-20% evolve with confirmed pulmonary embolism (PE) (11) with a mortality rate of 2-9% in these patients(5,6).

**Table 1.** Matching characteristics of cases *versus* controls.

| Matching variables | N (%) |  | p |
| --- | --- | --- | --- |
|  | CASES<br>(22) | CONTROLS (76) |  |
| **Sex** |  |  |  |
| Male | 11 (50%) | 39 (51.31%) | 0.911 |
| Female | 11(50%) | 37 (48.68%) |  |
| **Age at admission** |  |  |  |
| Median (IQR) | 7.85 (7.76) | 5.26 (7.68) | 0.133 |
| 0-1 y | 3 (13.63%) | 15 (19.73%) | 0.636 |
| 1-5 y | 6 (27.27%) | 27 (35.5%) |  |
| 6-10 y | 7 (31.81%) | 21 (27.63%) |  |
| 11-15 y | 6 (27.27%) | 13 (17.10%) |  |
| **Unit of admission** |  |  |  |
| Intensive Care Unit | 7 (31.81%) | 21 (27.63%) | 0.708 |
| Emergency Room | 0 | 2 (2.63%) |  |
| Nursery | 15 (68.18%) | 53 (69.73%) |  |

\*IQR, interquartile range

**Table 2.** Univariate analysis to identify potential risk factors for HA-VTE among cases *versus* controls.

| Risk factors for HA-VTE | N (%) |  | OR | *p*-Value |
| --- | --- | --- | --- | --- |
|  | CASES<br>(24) | CONTROLS<br>(76) |  |  |
| **Length of stay (days)** |  |  |  |  |
| Median (IQR) | 39 (39) | 6.5 (11) | 1.054 (1.018-1.092) | **0.003** |
| **More than 7 days** |  |  |  |  |
| **Yes** | 21 | 34 | 25.94 (3.32-202.81) | **<0.001** |
| **No** | 1 | 42 |  |  |
| **Nutritional status** | N=22 | N=73 |  |  |
| Obese | 5 (22.72%) | 9 (11.84%) | 2.19 (0.65-7.34) | 0.198 |
| Non-obese | 17 (77.27%) | 67 (88.15%) |  |  |
| **Surgery** |  |  |  |  |
| Yes | 5 (22.72%) | 23 (30.26%) | 0.678 (0.223-2.058) | 0.491 |
| No | 17 (77.27%) | 53 (69.74%) |  |  |
| **Immobility** |  |  |  |  |
| Yes | 13 (59.09%) | 38 (50%) | 1.444 (0.552-3.778) | 0.452 |
| No | 9 (40.90%) | 38 (50%) |  |  |
| **ICU admissions last 30 days** |  |  |  |  |
| Yes | 13 (59.09%) | 25 (32.89%) | 2.947 (1.111-7.815) | **0.003** |
| No | 9 (40.90%) | 51 (67.11%) |  |  |
| **Length of stay in ICU** |  |  |  |  |
| Median (IQR) | 8.5 (55) | 5.5 (5) | 1.059 (0.98-1.43) | **0.147** |
| **Mechanical ventilation** |  |  |  |  |
| Yes | 6 (27.27%) | 19 (25.00%) | 1.125 (0.385-3.288) | 0.829 |
| No | 16 (72.72%) | 57 (75.00%) |  |  |
| **Length of mechanical ventilation** |  |  |  |  |
| Median (IQR) | 10.5 (23) | 4 (8) | 1.201 (0.880-1.638) | 0.249 |
| **Infection** |  |  |  |  |
| Yes | 17 (77.27%) | 33 (43.42%) | 4.430 (1.481-13.249) | **0.005** |
| No | 5 (22.72%) | 43 (56.58%) |  |  |
| **Catheter** |  |  |  |  |
| Yes | 18 (81.81%) | 35 (46.05%) | 5.271 (1.630-17.045) | **0.003** |
| No | 4 (18.18%) | 41 (53.95%) |  |  |
| **L-asparaginase** |  |  |  |  |
| Yes | 6 (27.27%) | 2 (2.63%) | 13.875 (2.562-75.128) | **<0.001** |
| No | 16 (72.72%) | 74 (97.37%) |  |  |
| **Corticosteroids** |  |  |  |  |
| Yes | 14 (63.63%) | 32 (42.11%) | 2.406 (0.902-6.416) | 0.074 |
| No | 8 (36.36%) | 44 (57.89%) |  |  |
| **Heart failure** |  |  |  |  |
| Yes | 3 (13.63%) | 1 (1.32%) | 11.842 (1.165-120.32) | **0.010** |
| No | 19 (86.36%) | 75 (98.68%) |  |  |
| **Inflammatory/**<br>**autoimmune disease** |  |  |  |  |
| Yes | 3 (13.63%) | 6 (7.89%) | 1.842 (0.421-8.058) | 0.411 |
| No | 19 (86.36%) | 70 (92.11%) |  |  |
| **Liver failure** |  |  |  |  |
| Yes | 3 (13.63%) | 2 (2.63%) | 5.842 (0.910-37.486) | **0.038** |
| No | 19 (86.36%) | 74 (97.37%) |  |  |
| **Nephrotic syndrome** |  |  |  |  |
| Yes | 5 (22.72%) | 1 (1.32%) | 22.059 (2.418-201.219) | **<0.001** |
| No | 17 (77.27%) | 75 (98.68%) |  |  |
| **Neoplasia** |  |  |  |  |
| Yes | 6 (27.27%) | 15 (19.74%) | 1.525 (0.510-4.559) | 0.448 |
| No | 16 (72.72%) | 61 (80.26%) |  |  |

IQR, interquartile range

Hospital-acquired venous thromboembolism (HA-VTE) comprises multiple risk factors that should not be evaluated separately due to collinearity and multiple cause and effect relationships(7). For this reason, novel methods that allow a visual representation of theoretical assumptions of a process understudy that is easily interpreted by the reader, encouraging an iterative process of updating and revising theories about causal relationships, helping researchers in avoiding unintentional confounds and colliders (a common effect of two variables (i.e., exposure and an outcome) could be useful. Thus, the present study aims to improve the identification of risk factors for the development of HA-VTE in children using a novel approach, the directed acyclic graph (DAG), that seems to gather these necessary features(8) to deal with confounding factors of real-life.

## Design and Methods

### Study design

We conducted an observational, case-control study to evaluate risk factors for the development of hospital-acquired venous thromboembolism (HA-VTE) in children.

### Ethics and IRB approval

The study was approved by the Institutional Review Board (CAAE 58056516.0.0000.5264).

### Subjects

Children (newborn to 16 years old) hospitalized from May 24^th^, 2012 to August 31^st^, 2018, who developed hospital-acquired venous thromboembolism (cases) and randomized controls on a ratio of 1:3, matched by age (range), sex, unit of admission, and period of hospitalization (up to a 6-month interval). The study was held at a university hospital in Rio de Janeiro, Brazil (Instituto de Puericultura e Pediatria Martagão Gesteira), a 65-bed, tertiary care hospital (46 nursery beds, eight intensive care unit, and 11 emergency beds), with 25 pediatric subspecialties.

### Sample size

For the case-control study, 22 cases and 76 controls were selected on a ratio of 1:3 after statistical power calculation considering previous studies.

### Inclusion and selection criteria

Cases were defined as patients who developed hospital-acquired venous thromboembolism (HA-VTE) according to the International Society of Thrombosis and Hemostasis (ISTH)(9) criteria set as i) signs, symptoms or radiological diagnosis of venous thromboembolism (VTE): deep venous thrombosis and/or pulmonary embolism no sooner than two days of hospital admission; ii) Absence of documentation of signs or symptoms consistent with VTE in the admission history and physical exam; iii) Length of hospitalization of at least two days; iv) Diagnosis of VTE less than two days of hospital admission but a previous history of hospitalization in the last 30 days; v) Cases were classified as HA-VTE radiologically proved. Radiological diagnosis of VTE was confirmed using compression ultrasonography with Doppler imaging for objective confirmation of extremity DVT, with computed tomography (CT) or magnetic resonance imaging (MRI) for suspected extension into deep pelvic or abdominal veins, and spiral CT for pulmonary embolism (PE) confirmation. The Doppler ultrasound exam was registered in a separate file and not always was attached to the patient chart, so we revised it separately. The reason we created a classification of likely cases of VTE was that diagnostic modalities are not broadly available in our hospital. Cases were selected using four different strategies: i) Pharmacy software (disposal of anticoagulants); ii) International Classification of Diseases (ICD-10) codes for VTE; iii) Radiology report; iv) Active screening by medical students (only for the prospective phase of the study for one year, from August 1st, 2017 to August 31st, 2018. Controls were defined as hospitalized patients (for more than two days) and lack of history, signs, or symptoms of thrombotic events after evaluating patients charts and radiological records. Each case had an identification number and three controls were selected using our institution software database according to the following matching characteristics: i) Age group (0-1y; 1-5y; 6-10y; 11-15y); ii) Sex (male; female); iii) Unit of admission (Emergency, Nursery, and Pediatric Intensive Care Unit); iv) Period of hospitalization (from the onset of signs, symptoms or radiological diagnosis of VTE in the cases, what came first, to 180 days after discharge). If more than three controls were found, web-based randomization was performed (http://www.random.org). Cases and controls criteria were confirmed by two independent reviewers (MP and LRC).

### Exclusion criteria

Cases that only had thrombotic events other than VTE (deep venous thrombosis and/or pulmonary embolism) such as isolated superficial venous thrombosis, isolated thrombotic microangiopathy, and isolated arterial thrombosis were excluded from the study.

### Missing data criteria

Patients (cases or controls) that did not have adequate chart information during data collection were excluded.

### Data collection

Variables were categorized in baseline, follow-up, and outcome. Baseline variables evaluated in cases were case selection strategy (pharmacy, ICD-10, radiology report, or active screening), case thrombotic event (DVT, PE, superficial venous thrombosis, arterial thrombosis, and thrombotic microangiopathy), vessel involved in VTE by radiological reports (upper and lower limbs, pulmonary, intra-abdominal and central nervous system). Variables evaluated in both cases and controls included: demographic variables (age, gender, weight, height, body mass index (BMI), date of VTE diagnosis, length of stay, hospital unit location, and comorbidity using ICD-10 codes), clinical data on VTE prophylaxis (heparin, vitamin K antagonists, pneumatic compression devices, early mobilization, and physiotherapy), putative risk factors (prematurity, obesity according to Centers for Disease Control and Prevention (CDC) definition(10,11), surgery, immobilization for more than 72 hours, reason for immobilization (critical patient, post-operative, physical disability, plaster), ICU admission, length of stay in ICU, mechanical ventilation, days on mechanical ventilation, trauma, trauma on the site of VTE, central venous catheterization (information regarding type, location, unsuccessful previous attempt), drugs (L-asparaginase and corticosteroids), heart failure, inflammatory/autoimmune disease, liver failure, nephrotic syndrome, thrombophilia, family history of thrombosis, dehydration, polycythemia, and cancer. Follow-up variables for cases were the type of VTE therapy (heparin, vitamin K antagonist, vena cava filter, thrombolysis), the dosage of anticoagulant used, bleeding and its classification (according to ISTH(12)), and laboratory test used for anticoagulation monitoring: international normalized ratio (INR), Partial Thromboplastin Time (TTP), and anti-factor Xa. Outcome variables were death (cases and controls), pulmonary embolism (for patients with previous DVT), the extension of the VTE (radiology proved), and the transformation from non-occlusive to occlusive VTE.

### Statistical analysis

Descriptive statistics were used to define distributions of continuous variables, frequencies, and proportions of categorical variables, which were then compared between cases and controls using Mann-Whitney U and χ^2^ tests, respectively. Fisher’s exact test was used in place of χ^2^ in instances of a two-by-two table with a frequency value of 5 or under in at least one cell. Results were expressed as odds ratios (ORs), with accompanying 95% confidence intervals (95% CIs) calculated by the Wald method.

To evaluate the trend rate of HA-VTE incidence in the last years, we implemented a temporal series analysis with data from 2012 to 2017. The Autoregressive Integrated Moving Average (ARIMA) was adopted. The cases were divided into three different age categories: all ages, less than six years, and from 6 to 15 years old. The linear correlation between the HA-VTE rate and the years use evaluated by R-square (coefficient of determination).

Due to a large number of potential variables that could confound or mediate the causal effect of the exposure (risk factors of HA-VTE) on the outcome (VTE), a directed acyclic graph (DAG) model was created to reduce bias, improve transparency, and increase precision on the analysis.

Variables included in DAG were clustered in four different groups: (1) exposure variables (mechanical ventilation, liver failure, ICU admission, catheter, heart failure, obesity, and length of stay), (2) forced-variables (heart failure, obesity, and liver failure – not linked to other risk factors in the model), (3) non-forced variables (immobilization, local trauma, nephrotic syndrome, surgery, infection, hematologic malignancies, L-asparaginase, autoimmune/inflammatory disease, corticosteroids, and venous thromboembolism) and (4) non-observed variables (dehydration, first-degree family history of thrombosis, and thrombophilia – variables with missing data not included in the statistical analysis). We selected eight main exposure variables: central venous catheter, intensive care unit stay, mechanical ventilation, and length of stay were based on the systematic review and meta-analysis of risk factors, and risk-assessment models published by Mahajerin A *et* al. (7). Other risk factors comprised in DAG such as liver failure, heart failure, immobilization, local trauma, nephrotic syndrome, surgery, infection, hematologic malignancies, L-asparaginase, autoimmune/inflammatory disease, and corticosteroids were also included based on our assumption that diseases work as leading players on the development of VTE and are usually connected with other risk factors. One example is that we typically do not see children with infection alone having VTE. Obesity was also included because it was one of our hypotheses as a risk factor for pediatric VTE, and it is not yet fully explored in other studies. After selecting the exposure variables, we created directed paths using a browser-based software DAGitty (http://www.dagitty.net) (Figure 1). Variables were represented according to the software pattern (Figure 2). A family history of thrombosis and thrombophilia are described as unobserved variables because we intended to collect it, but essential information was lacking during data analysis.

We used the algorithm proposed by Greenland *et* al. (13–16) to control potential confounders in the context of rare outcomes, data sparsity, and multicollinearity. It is especially useful when the number of covariates is significant to the study size. Therefore, we implemented the following steps: (1) selection of the variables that are appropriate to include using a causal directed acyclic graph (DAG) to exhibit theorized causal relations among variables; (2) division of the variables into three classes: (i) the main exposure X; (ii) forced-in variables (e.g., age, sex) (iii) the non-forced variables which will be candidates for deletion; (3) building a Full Model, through conditional logistic regression model, including all main exposure terms, forced-in variables, and non-forced variables. The next step was to select which variables of the Full Model would be included in the final parsimonious adjusted model (Final Model). We applied three methods to obtain this Final Model to validate the results of each modeling process: (1) assess, and thereby minimize, mean squared error (MSE) using forward selection strategy, as we indicated sparse-data bias; (2) usual forward method, using the likelihood ratio stepwise selection from the variables presented in the Full Model; (3) usual backward deletion method, using likelihood ratio stepwise selection with a previous traditional multicollinearity analysis from the variables presented in the Full Model. The first method was implemented in Microsoft Excel® (2016) software, and the two last traditional methods, using SPSS Statistics® (version 25.0. Armonk, NY: IBM Corp.) Interestingly, all methods resulted in the same Final Model (table 3).

**Table 3.** Multivariate regression models. Statistical significant differences are in bold.

|  | FINAL MODEL |  |
| --- | --- | --- |
|  | OR<br>(95%CI) | p |
| **Length of stay** | **1.108** | **0.011** |
|  | **(1.024-1.199)** |  |
| **L-asparaginase** | **27.184** | **0.021** |
|  | **(1.639-450.982)** |  |
| **Nephrotic syndrome** | **31.481** | **0.039** |
|  | **(1.182-838.706)** |  |
| Mechanical ventilation | 0.18<br>(0-1.318) | 0.067 |

C.I: confidence interval; OR: odds-ratio

## Results

Given the 6,295 hospitalizations during six years of the study, this yielded an average HA-VTE of 6.25 per 10,000 hospitalized children per year. Twenty-six cases of hospital-associated venous thromboembolism (HA-VTE) were diagnosed during the six years of observation but only 22 cases were enrolled after application of the validation criteria for venous thromboembolism (VTE) cases selection and loss due to missing data (Figure 3).

Most cases (86.7%; n=19/22) were found using data bank of the pharmacy sector stored in MV 2000® software (Recife, Pernambuco, Brazil). The twenty-two cases were matched to 76 randomly selected controls. There was no difference in the frequency of VTE between cases and controls regarding sex, age, and unit of admission as expected due to matching (Table 1). We did not observe peak VTE in infancy due to the absence of the Neonatal Intensive Care Unit and Adolescent ward in the hospital.

We found a significantly increased trend rate of HA-VTE throughout the years in our institution (Figure 4_)_, which was more pronounced in the <6 years age group.

Four patients (4/22; 18.2%) presented with signs and symptoms of VTE less than 48 hours of admission and were previously hospitalized in the last 30 days. Regarding the VTE site, two patients (2/22; 9.1%) had a pulmonary embolism (PE), including one associated with deep venous thrombosis (DVT). One patient (1/22; 4.5%) had superficial venous thrombosis (SVT) associated with DVT, and two patients (2/22; 9%) had thrombotic microangiopathy (TMA) associated with DVT.

The median time from hospital admission to VTE diagnosis was 28.5 days (IQR 32 days) and the most frequently associated signs and symptoms of DVT were edema (19/22; 86.3%), pain (6/22; 27.3%), discoloration of the limb (5/22; 22.3%) and warmness (2/22; 9.1%). Two patients (2/22; 9.1%) had PE and presented with the following symptoms: one with cough and hemoptysis and the other with dyspnea, tachypnea, cough, and tachycardia. Only one patient had cerebral venous sinus thrombosis (CVST) and presented with seizures. He had a diagnosis of acute lymphoblastic leukemia (ALL) and was treated with L-asparaginase. DVT cases were diagnosed using Doppler ultrasound and PE with a CT scan. The 22 patients had 38 sites of VTE which included lower limbs (26/38; 68.4%); intra-abdominal (4/38; 10.5%) – inferior vena cava (2/4; 50%), hepatic vein (1/4; 25%) and renal vein thrombosis(1/4; 25%); central nervous system (3/38; 7.9%), upper limbs (3/38; 7.9%) and pulmonary arteries (2/38; 5.3%).

Only one patient (1/22; 4.5%) developed VTE while on the prophylactic use of subcutaneous low molecular weight heparin (LMWH) in a dosage of 1 mg per kg, once daily, on day 30. She was a 15-years old adolescent with encephalitis admitted in the ICU with several risk factors, including immobilization (30 days), mechanical ventilation (14 days), and catheter on the same site of VTE (right common femoral vein). Another patient developed recurrent VTE even on therapeutic LMWH. She was a 3-year old patient with Down syndrome with several risk factors, including high-risk ALL treated with L-asparaginase and corticosteroids, gastrointestinal infection, febrile neutropenia, obesity, and thrombophilia (combined heterozygosity for methylenetetrahydrofolate reductase and factor V Leiden). DVT occurred on the left femoral common vein with previous attempting of catheter insertion.

The risk factors for HA-VTE among cases *versus* controls are presented in Table 2. The median length of stay was higher in cases (39 days – IQR: 39) than controls (6.5 days – IQR: 11). Central venous catheter (CVC) use was higher in cases (81.81%) than controls (46.05%). Long-term CVCs were identified in 16.66% of catheterized patients, with the remainder (83.33%) having short-term CVCs (Peripherally Inserted Central Catheter [PICC] lines, temporary jugular, subclavian or femoral lines). ICU admission in the last 30 days was seen more frequently in cases (13/22; 59.09%) than controls (25/76; 32.89%). Infection was also more frequent in cases (17/22; 77.27%) than controls (33/76; 43.42%) with no difference between local or systemic ones. L-asparaginase use was higher in cases (6/22; 27.27%) than controls (2/76; 2.63%). Nephrotic syndrome was seen in 22.72% (5/22) of cases compared to 1.32% (1/76) of controls. Liver failure was more common in cases (3/22; 13.63%) than controls (2/76; 2.63%). We did not observe any difference between cases and controls regarding obesity, surgery, immobility, mechanical ventilation, length of mechanical ventilation, corticosteroid use, inflammation/autoimmune disease, and neoplasia.

For estimating the direct effect of the exposure variables (heart failure, ICU admission, length of stay, liver failure, mechanical ventilation, catheter, and obesity) on the outcome (venous thromboembolism), the software recommended doing an adjustment on corticosteroids, hematologic malignancies, immobilization, infection, L-asparaginase, local trauma, nephrotic syndrome, and surgery (Supplement Material S1 and S2). After that, we created two different multivariate regression models: (1) Full Model, utilizing all risk factors for VTE that were statistically significant, and a (2) Final Model after adjustment (Table 3). Length of stay (LOS), liver failure, L-asparaginase use, and nephrotic syndrome were independent risk factors for hospital-associated venous thromboembolism in children.

## Discussion

We found a significant increase in the incidence of HA-VTE across all age groups from 2012 to 2017, with a similar trend rate reported in a multicenter study in the United States (1).

To our knowledge, this is the first study of hospital-associated venous thromboembolism using a directed acyclic graph (DAG) to deal with multiple confounding risk factors.

The DAG is a visual representation of theoretical assumptions of a process understudy that is easily interpreted by the reader, encouraging an iterative process of updating and revising theories about causal relationships. A DAG-informed approach helps researchers in avoiding unintentional confounds and colliders (a common effect of two variables (i.e., exposure and an outcome), that is represented in DAG by the two arrows pointing toward it(13), arousing from these variables). The DAG approach has limitations because it does not eliminate all sources of bias; it is time-consuming, requires a broad review of the literature and evidence-based understanding of causal relationships. It is also a pre-requisite to choose between minimally enough adjustment sets considering all variables that are well measured and well specified with little missing data. Likewise, as with any analytic approach, DAGs cannot account for unmeasured confounding or potential covariates not determined from the literature(17). There is a robust literature supporting DAG theory and technique (8,13,18), but this approach has not yet been broadly adopted, although it has been highly recommended by experts in the field of biostatistics(15).

We found the **length of stay (LOS), L-asparaginase use**, and **nephrotic syndrome** to be independent risk factors for hospital-associated venous thromboembolism in children. In a recent systematic review and meta-analysis of risk factors and risk-assessment models by Mahajerin A et al. (7), a central venous catheter (CVC), intensive care unit stay, mechanical ventilation, and length of stay were identified as leading risk factors in the case-control studies. Most of these studies did not systematically evaluate the more common co-morbidities. Those factors, besides mechanical ventilation, were also considered risk factors (p<0.005) in univariate analysis in our study, but not all of them were maintained in the final multivariate model. This finding emphasizes the importance of considering the co-morbidity as essential covariable to be included in VTE case-controls studies as interventions (i.e., catheter) occur in sick children.

**Length of stay (LOS)** has been a consistently identified risk factor but the mechanism by which it increases the risk of VTE is not fully understood. It works as a proxy that also covers unknown risk factors not yet investigated. We postulate some theoretical relationships that were represented by DAG analysis (figure 1). Length of stay (LOS) is a risk factor that remains in many case-control and non-case-control studies, which suggests that it acts as a proxy for known and unknown risk factors. Among the known risk factors, some are very insightful, such as length of stay in the ICU, trauma, immobilization, and surgery. In a scenario of multiple risk factors with numerous medical conditions and interventions, it is expected that a meticulous structured statistical model is needed, a good reason for using the DAG concept. Following DAG analysis, LOS persisted as a critical risk factor, which suggests that there are relevant factors not yet considered in our theoretical model and, therefore, not controlled in multivariate analysis. One could argue that the persistence of LOS as a risk factor would be a small sample size issue. However, even larger studies (19,20), including a recent comparative validation study of risk assessment models for pediatric hospital-acquired venous using the Children’s Hospital-Acquired Thrombosis (CHAT) registry(21), still confirm the importance of LOS. Another reason would be the fact that unknown elements, such as biological or biochemical factors involved in the genesis of VTE, are not yet understood, identified, or available in clinical studies. For this reason, we still do not have enough knowledge to explain all the relationships related to LOS and venous thromboembolism, an issue that requires further investigation.

Acute lymphoblastic leukemia (ALL) is the most common type of pediatric cancer and, therefore, the most frequent malignancy associated with VTE in childhood. The incidence of VTE ranges from 5 to 70% when imaging studies identify asymptomatic cases. **Asparaginase** is a crucial chemotherapy agent in the treatment of ALL and can induce an acquired deficiency of protein C, S, and antithrombin(22). Most thrombotic events occur in the upper venous system and the central nervous system (CVST)(23) with a potential of severe consequences. Adult studies showed that prophylactic anticoagulation could be safely administered without increasing the number or severity of bleeding events, resulting in a reduction in the cumulative incidence of VTE(24) in ALL patients. Hence, VTE prophylaxis(25) or protein replacement with fresh frozen plasma(26) has recently been proposed in patients with ALL during the induction period with pediatric clinical trials ongoing. In our study, prophylactic anticoagulation in ALL was not employed.

**Nephrotic syndrome** is another prototype for pediatric VTE with multifactorial and complex pathophysiology. The nonselective proteinuria results in hemostatic abnormalities mainly due to loss of anticoagulant and fibrinolytic proteins (such as antithrombin III, plasminogen, protein S, and plasmin) and increase of prothrombotic substances (factor V, VIII, alpha-2 macroglobulin, thromboxane A2, and fibrinogen) produced by the liver to balance the loss of anticoagulant elements contributing to a procoaguable state(29). Other reported risk factors for venous thromboembolism (VTE) in this population include infection, CVCs, diuretics, and intravascular volume depletion(30). In our study, 5 of 6 (83.33%) patients with nephrotic syndrome developed VTE. Infection (60%), CVC (40%), and ICU admission (20%) were also seen in most patients. Our study showed a higher percentage (83.33%) of nephrotic patients with VTE, considering a recent cohort study of 370 children with nephrotic syndrome admitted to 17 pediatric hospitals across North America from 2010 to 2012, where only 11 (3%) of patients developed VTE(30).

Mechanical ventilation was not statistically significant in the final model but remained to better adjust confounding factors.

The limitations of our study include the small sample size (as a result of single-center research), the retrospective nature and, the missing data of some patients, i.e., lack of data on hypercoagulability, absence of radiological documentation of all VTE events - a real-life situation in retrospective studies in underdeveloped countries. One of the reasons our sample was not larger was the fact our hospital does not admit patients with solid malignancies, does not perform cardiothoracic and orthopedic surgery and, the emergency department is referred only for patients that are attended in the outpatient clinic (i.e., not trauma patients). There are some explanations about why the findings in our study were different from other publications. Potential covariates, unmeasured confounding, and lower statistical power could be essential reasons that justify these differences. On the other hand, we evaluated the co-morbidities in our DAG model, and this could have lessened the significance of medical interventions as the leading risk factor of HA-VTE. We hope that the more extensive ongoing studies that will probably consider the role of co-morbidities can establish the real weight of the medical intervention in HA-VTE in children.

Our study has some important strengths. To our knowledge, this is the first case-control study of pediatric HA-VTE using DAG analysis worldwide, and the first pediatric case-control HA-VTE study in Brazil. The DAG-based approach in our study highlighted the importance of medical conditions, appeared as the most critical variables, and made us question if therapeutic interventions would be only triggers or proxies (“tip of the iceberg”) for the observed thrombosis. Researchers should consider medical conditions (i.e., nephrotic syndrome, ALL) in risk-assessment models besides directing spotlights to interventions (i.e., catheters). We reported the importance of medical conditions on the genesis of HA-VTE using a DAG-based approach, which makes it possible to clarify the influence of confounders and multiple causalities, such as catheter, a significant risk factor highlighted in several studies. This approach is rational since children that use intravenous devices are sick, so it is not reasonable to put all the weight on a catheter since other risk factors play an essential role in VTE. Besides that, we reported an increase of HA-VTE events along the years of the study, which suggests that educational actions should be performed to prepare the clinical team not only to deal with these patients but to teach medical students and residents since it is a university hospital. Finally, an international effort should be made to increase the robustness of data and to balance the different characteristics of pediatric hospitals in the globe to create offset risk-assessment models and, therefore, adequate protocols for HA-VTE prophylaxis in children.

## Data Availability

All data is included in the article and Supplemental material.

## Supporting information

**S1 Table. DAG flowchart code**.

**S2 Table. Conditional independence and minimal sufficient adjustment proposed by the DAG model**.

## Authors’ contributions

LRC, MP, FRS, ESC, LRB, and MGPL contributed to the conception and design of study. LRC and MP contributed to acquisition of data. LRC and MGPL contributed to analysis of data and with the interpretations of data. LRC, FRS, ESC, LRB, and MGPL were the major contributors in writing the manuscript. All authors revised the manuscript critically for intellectual content. All authors read and approved the final manuscript and the listing of authors.

### Ethics approval and consent to participate

The study was approved by the Institutional Review Board (CAAE 58056516.0.0000.5264).

## Acknowledgments

We acknowledge “TROMBOPED” medical students of Universidade Federal do Rio de Janeiro for the participation on the data collection.

---

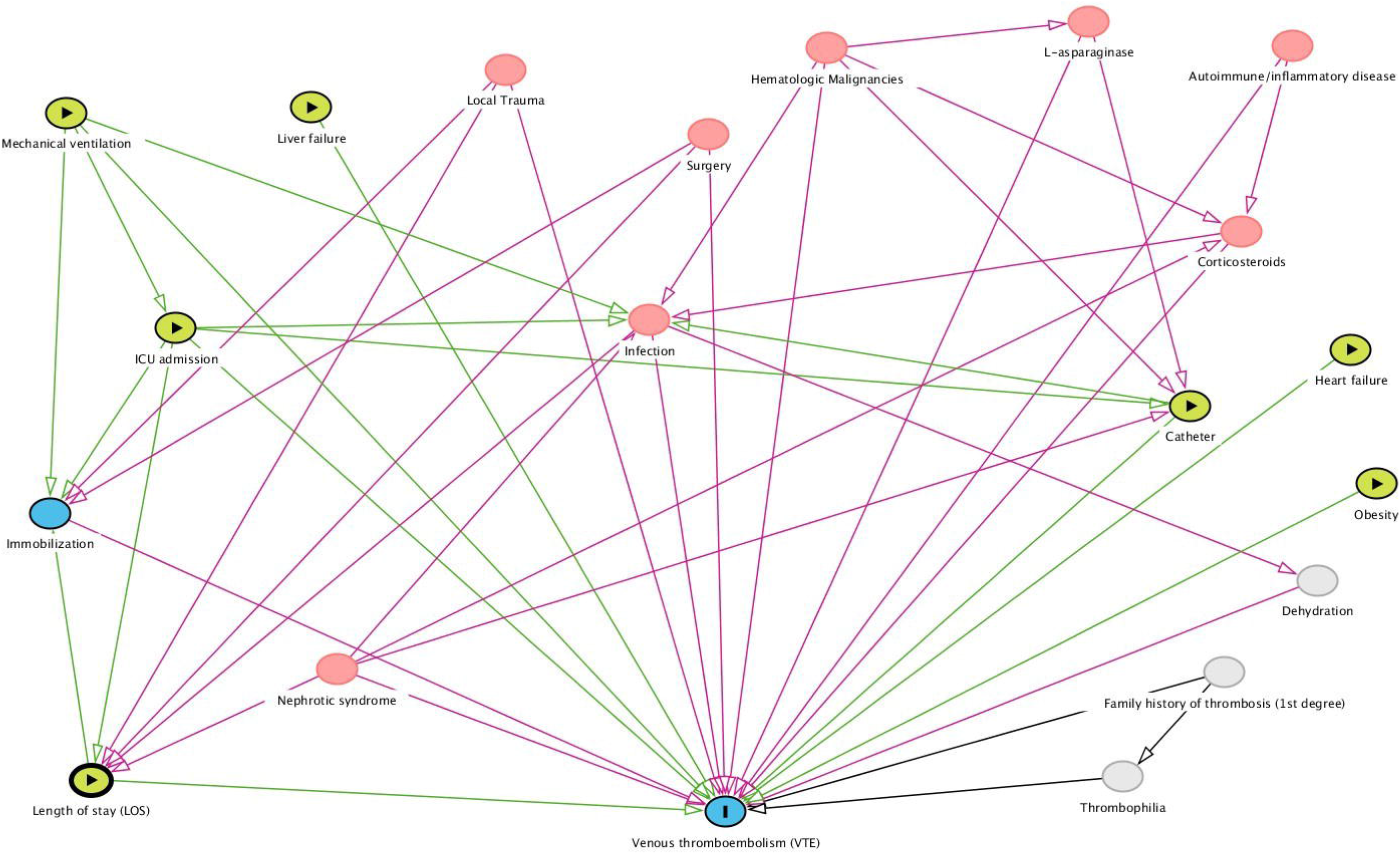

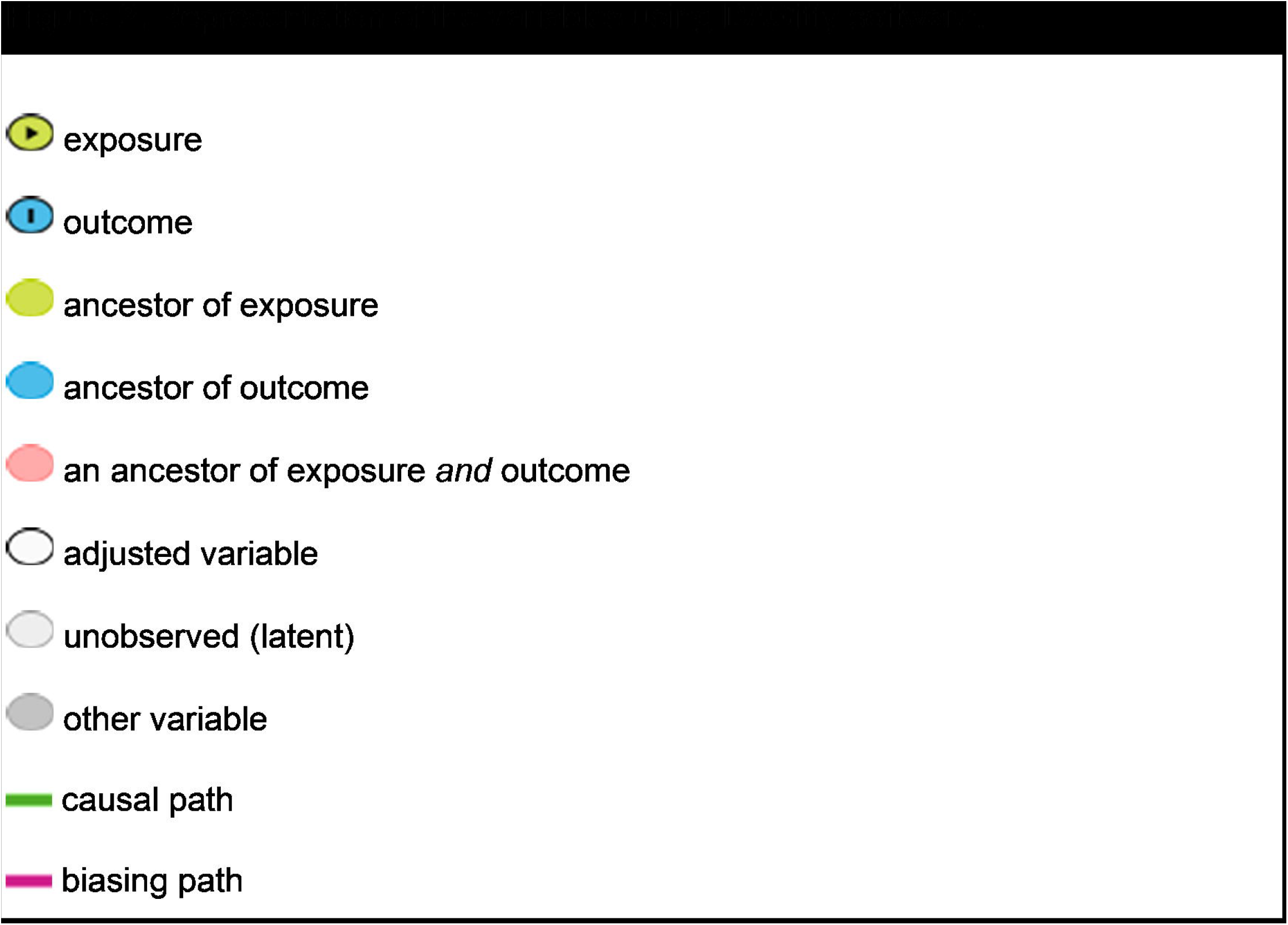

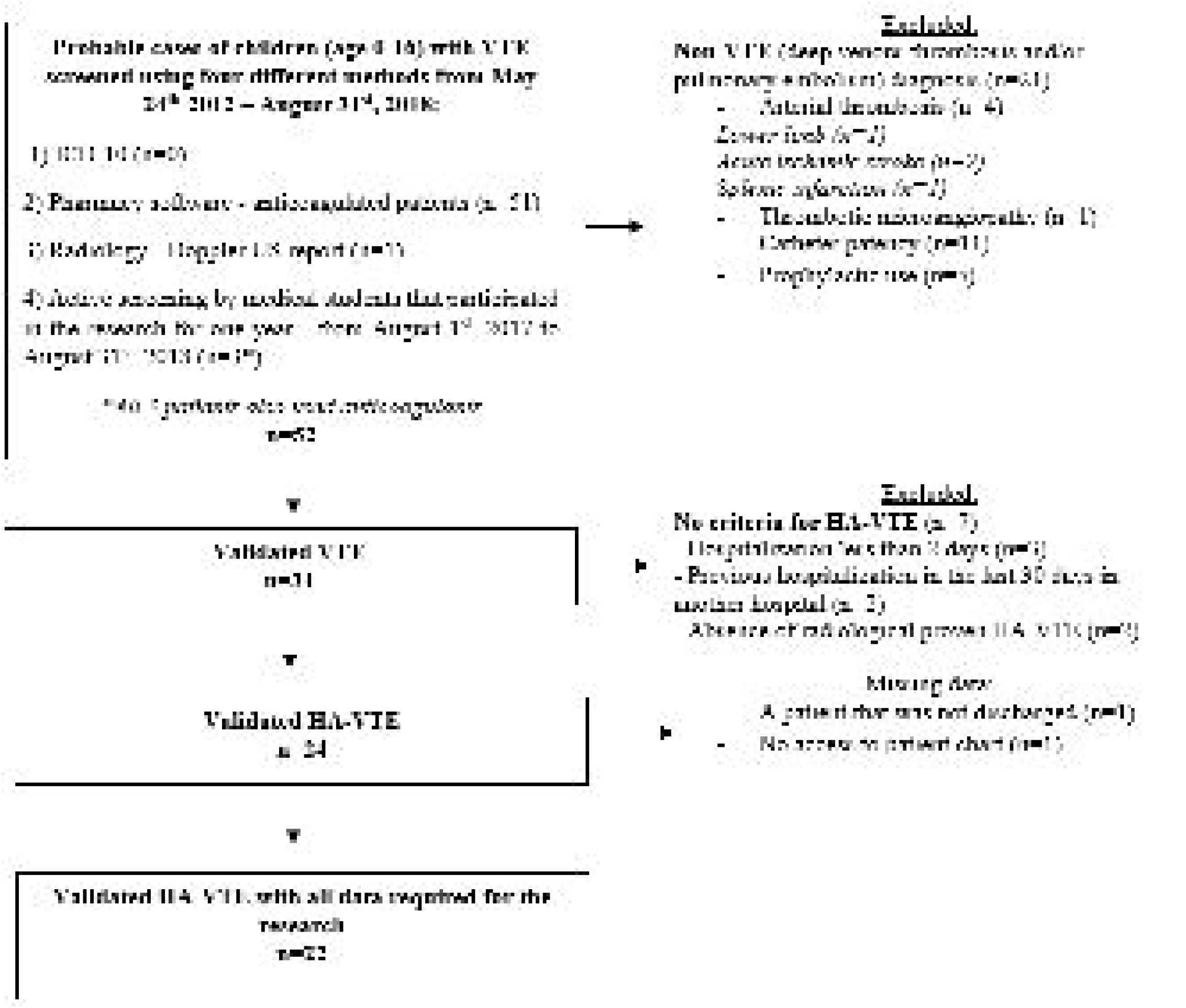

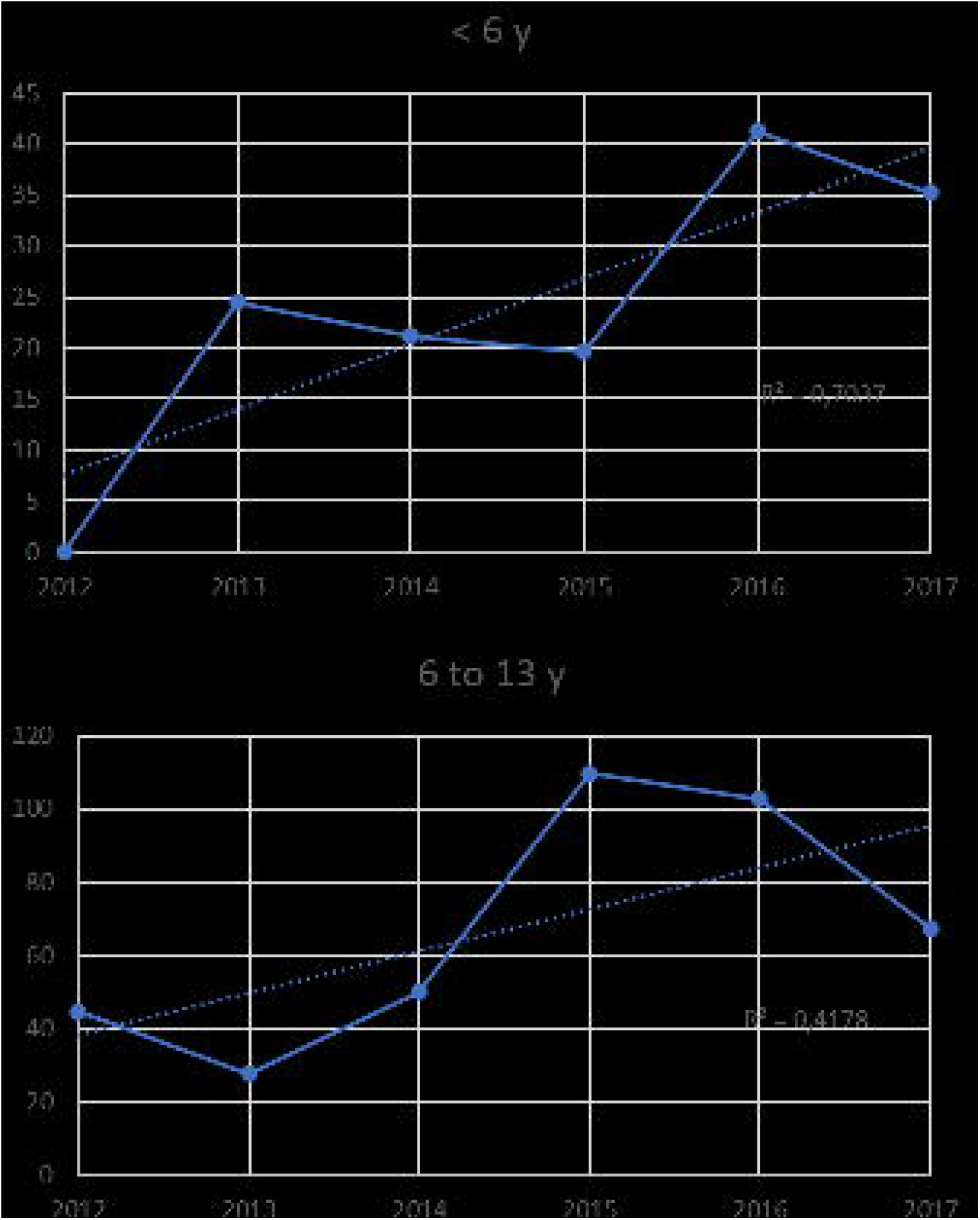

